# Analyzing the Mechanism Behind Age-agnostic Prediction of Diastolic Dysfunction Using Echocardiography Variables in Deep Neural Networks

**DOI:** 10.1101/2023.09.17.23295672

**Authors:** Ankush D. Jamthikar, Rohan Shah, Marton Tokodi, Partho P. Sengupta, Naveena Yanamala

**Affiliations:** Division of Cardiology and Hypertension, Department of Medicine, Rutgers Robert Wood Johnson Medical School, New Brunswick, NJ, USA; Heart and Vascular Center, Semmelweis University, Budapest, Hungary

**Keywords:** deep neural network, echocardiography, left ventricular diastolic dysfunction, age, sex prediction

## Abstract

**Objective:** This investigation delved into the inner workings of previously published Deep Neural Networks (DNNs) designed to detect changes in diastolic function related to age using echocardiographic parameters. The primary goal was to decipher the predictive mechanism and determine whether biological age and gender played concealed roles within the DNN model.

**Methods:** We conducted a secondary analysis of a previously published DNN model that was trained using data from 1,009 patients (average age: 62±17 years, 57% females) to forecast risk phenogroups based on nine echocardiographic parameters. This model was assessed on both an internal cohort (n=243, mean age = 62 ± 17 years, ∼57% females) and an external validation cohort (n=5596, mean age = 76 ± 5 years, ∼57% females). To forecast biological age and gender, we developed linear regression and classification models employing hidden layer activations from the DNN. Model performance was assessed using Pearson’s correlation for regression, accuracy, and area under the curve (AUC) metrics for classification.

**Results:** Upon scrutinizing the hidden layer activations, we observed that the model accurately captured biological age in both younger and older populations, particularly in low-risk phenogroups, with robust correlations for the entire population (0.94, p<0.001), males (0.90, p<0.001), and females (0.94, p<0.001). In high-risk phenogroups, the correlations were lower, standing at 0.31 (p=0.274) for the entire population, 0.76 (p=0.003) for males, and 0.11 (p=0.723) for females. Predicting gender as an underlying factor resulted in an accuracy rate of 58.02% and 52.27%, accompanied by an AUC of 0.65 for both validation cohorts.

**Conclusion:** This study underscores that the latent space within DNNs maintains a link with age in relation to diastolic functional parameters, offering a solution that is independent of age for predicting diastolic dysfunction. The dissection of the network can further enhance our comprehension of the information learned by DNNs, thereby providing novel pathophysiological insights for medical professionals.

## Introduction

Neural network dissection is a methodology that deciphers the intricacies of deep neural networks, often seen as “black boxes” due to the difficulty in understanding their operation ^1 2^. This technique illuminates the network’s functional behavior by examining individual neurons, hidden layers, and their connections systematically ^1 3^. Furthermore, it enables the exploration of learned representations in the network, thereby identifying specific features or concepts prioritized during decision-making or classifications ^2 4^. The derived insights clarify the network’s behaviour and facilitate model optimization, interpretability, and domain-specific understanding. It finds applications across diverse domains like computer vision, natural language processing, and biomedical research ^2 3 5–8^.

In our prior research, we developed a deep-learning model for the echocardiographic evaluation of left ventricular diastolic dysfunction (LVDD) in younger cohorts ^9–11^. LVDD is a condition where the left side of the heart struggles with relaxation and blood filling, affecting its pumping efficiency ^12^. Aging significantly contributes to structural and functional changes in the heart, and these alterations have been linked to LVDD. Despite not incorporating age adjustment in the training phase, our deep learning-based model successfully differentiated between low-risk and high-risk phenogroups across younger and older cohorts.

Building on our previous model’s age-independent performance, our current study explores how the model learns the age-related association with left ventricular functional parameters. We hypothesize that the model developed for predicting low and high-risk phenogroups of LVDD can also identify age or sex-related changes. To delve into our deep learning models’ age-or sex-related learning, we dissected the neural network model and examined its latent space of trained parameters and hidden neural activations.

## Methods

### Study Population

This study includes two previously published databases that comprises a local derivation cohort and an external validation cohort ^9^. The local derivation cohort included 1,252 patients (mean age = 62±17 years, ∼57% females), out of which 799 participants (mean age = 66±17 years) were retrospectively enrolled from the Icahn School of Medicine at Mount Sinai, New York ^9 10^. The remaining 453 participants (mean age = 56±17 years) were included in two prospective trials at the West Virginia University School of Medicine. Our previous studies have provided a detailed study protocol and inclusion and exclusion criteria ^9–11^. The external validation cohort includes the 5,596 participants from the Atherosclerosis Risk in Communities (ARIC) analysis, a prospective epidemiologic cohort study started in 1987 to investigate the etiology of atherosclerosis and its clinical sequelae. The ARIC study enrolled 15,792 participants aged 45-64 from four US communities ^13^. The current study analyzed 5,596 participants (mean age=76±5 years) who underwent echocardiographic assessment during the fifth visit between 2011 and 2013. Our previous studies have discussed the details of the inclusion and exclusion criteria and the information about the institutional review board approval ^9 11 13^. Note that the current study has a specific emphasis on network dissection, investigating the potential of hidden activations to capture age-associated changes in echocardiographic parameters. The unique design of this study sets it apart from our previously published studies, which concentrated on assessing left ventricular diastolic dysfunction (LVDD) in younger and elderly cohorts ^9–11^.

### The DeepNN model of LVDD

In our previous study ^10^, we described an unsupervised method that transforms the nine echocardiographic variables into a similarity network using topological data analysis (TDA), which forms a continuous loop based on the varying degree of systolic or diastolic dysfunction. The TDA-based similarity network assigns low-risk or high-risk labels to patients using their nine routinely measured echocardiographic variables such as ejection fraction, left ventricular mass index, early diastolic transmitral flow velocity, late diastolic transmitral flow velocity, E/A ratio, early diastolic relaxation velocity, E/e’ ratio, left atrial ventricular index, tricuspid regurgitation velocity. The brief information about these echocardiographic parameters along with their guideline-based thresholds for abnormality are provided in the supplementary material (Section A). After labeling the patients as high-risk or low-risk using the TDA-based approach, a cloud-based automated machine-learning platform (OptiML, BigML, Corvallis, Oregon) was used to train multiple machine learning algorithms and selected the best performing algorithm as the DeepNN model. Our group has already published the detailed methodologies for the TDA-based similarity network and the DeepNN model in ^9 10^.

### Development of an emulator model for investigating latent space information

The DeepNN model was explicitly developed for predicting the high-risk or low-risk phenogroups of LVDD using the commercial cloud-based BigML platform ^9^. Since it is not possible to extract hidden weights from the DeepNN model developed using the BigML framework, another similar multi-layered deep neural network (DNN) model was developed using an open-source Python-based library. This new DNN model emulates the characteristics of the existing DeepNN model (hence called emulator model hereafter) and provides access to trained weights that are useful to investigate the information about the latent space. The training of the emulator model is performed on the same local derivation cohort (n=1,252) with high-risk or low-risk patient clusters used as class labels. Out of 1252 patients, 80% (n=1009) are used for model training, and 20% (n=243) are used for internal validation. Figure 1 (A) shows the internal architecture of the emulator model that predicts the binary risk profiles from the nine echocardiography parameters. The model has four hidden layers, each with 13, 27, 63, and 6 neurons. The choice of the number of layers and the neurons in each layer is governed by the open-source Python-based Optuna-3.0.1 library for hyperparameter tuning ^14^. During the training process of the emulator model, responses from all the hidden neurons (aka ‘neural activations’) are recorded. The latent information within such neural activations is further used to investigate the association with the actual age or sex of a person.

**Figure 1.**
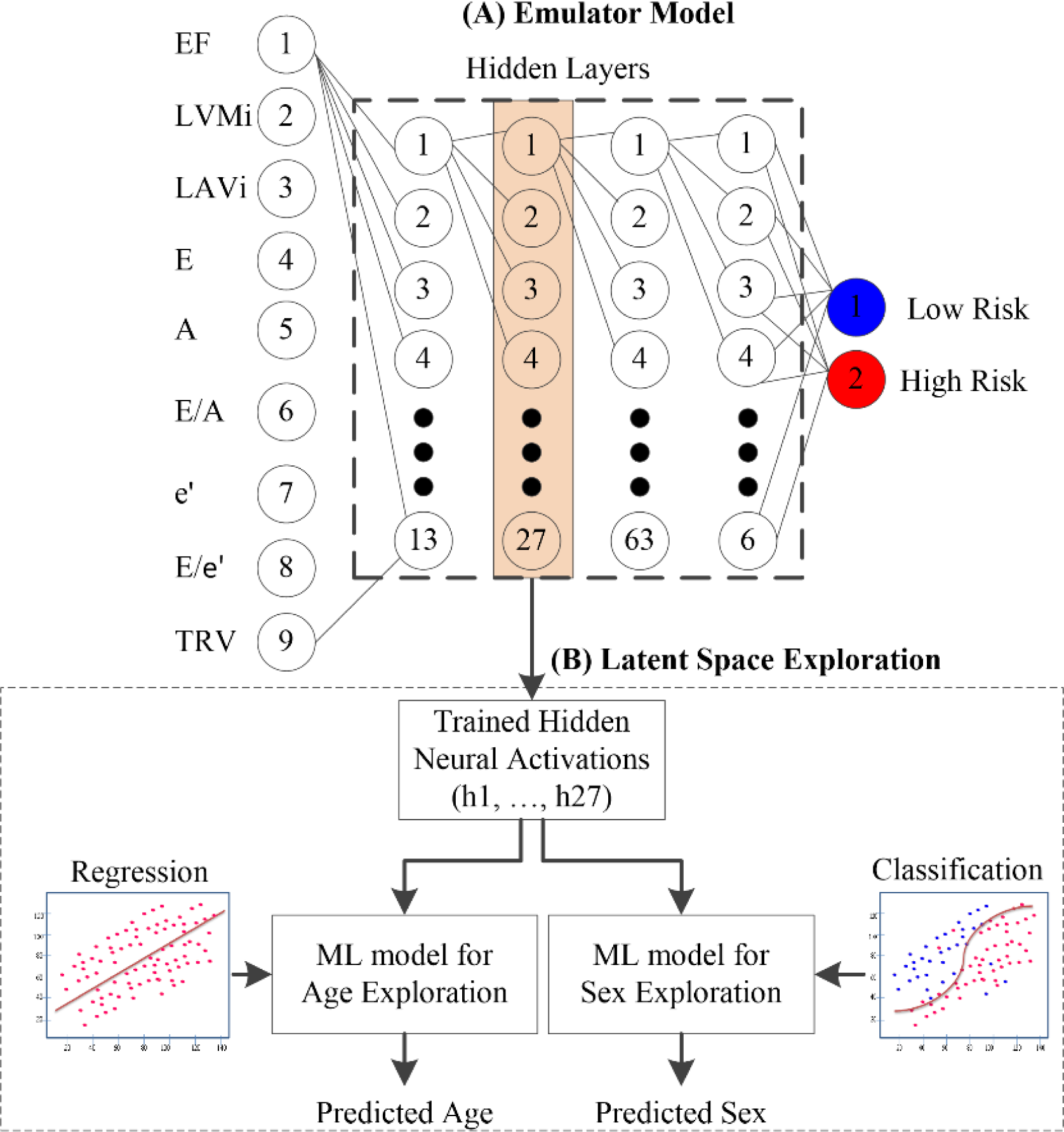
(A) The architecture of the emulator model predicting low- and high-risk potential phenogroups of LVDD and (B) latent variable exploration from the neural activations. EF – ejection fraction, LVMi – left ventricular mass index, LAVi – left atrial volume index, E – early mitral inflow velocity, A - late mitral inflow velocity, e’ – early diastolic mitral annular velocity, TRV – tricuspid regurgitation velocity.

### Latent space exploration for the emulator model

To investigate if the emulator model is age- and sex-agnostic, the hidden neural activations from each layer are used to train separate regression and classification models to predict actual age and sex, respectively (Figure 1B). For age prediction, a linear regression model is trained on the neural activations obtained from the training data (n=1009). Since there are four hidden layers with a different set of neurons, four independent linear regression models are trained, and the corresponding coefficients are recorded. The trained coefficients of the linear regression model are then used to transform the neural responses for the internal and external validation databases into the predicted biological age of patients. Note that the nine echocardiography parameters drive the predicted age and may differ from the actual age of patients. Linear regression is the most fundamental approach for finding the association between continuous variables and, thus, is adopted in this study to investigate the link between neural activations and actual age.

We also investigated the association between neural activations and sex as a latent variable. Since sex is a dichotomous variable, a classification model is trained on the neural response of the training database. Then this trained model is used to transform the neural response from the validations databases into the predicted sex of patients. Five popular machine learning-based classification algorithms, such as logistic regression ^15^, support vector machine ^16^, bagging ^17^, random forest ^18^, and extreme gradient boosting ^19^ are compared to determine the best choice for predicting sex as a latent variable from the neural responses.

### Performance Evaluation and Statistical Analysis

To verify the equivalence between the previously proposed DeepNN model ^9^ and the emulator model was established using (i) the five performance evaluation metrics such as sensitivity, specificity, F1-score, accuracy, and area under the curve (AUC), and (ii) the confusion matrix between the predicted labels from both models. Outlier detection using the inter-quartile range and missing value removal impacting ∼5% of the total dataset was performed on both the derivation and validation cohorts before the statistical analysis. In latent space exploration, the association between the actual age and the echocardiography-driven predicted biological age obtained is examined using Pearson’s correlation coefficient, i.e., the r metric. The combined internal and external validation data was analyzed using scatter plots. These plots compared the mean over a 5-year age range for both actual and biological age categories, depicting the mean ± standard error around the mean age. The two-tailed t-test is used to determine the statistical difference between the predicted age in the low-risk and high-risk categories with a level of significance, p<0.05. Patients were clustered into groups using hierarchical clustering based on a similarity measure ^20^. The association between hidden neural activations and sex is governed by the optimal classification algorithm, whose performance is evaluated using the six performance evaluation metrics discussed above.

## Results

### Baseline Characteristics

The baseline characteristics of the local derivation cohort and the external validation cohort are provided in Table 1 and Table 2. As shown in the tables, the actual age and sex, along with nine echocardiographic parameters, are significantly associated with the phenogroups of diastolic dysfunction. The average age in the derivation cohort is 62 ± 17 years (18 - 97 years), and in the external validation cohort is 76 ± 5 years (66 - 90 years). This age distribution shows that the derivation cohort mostly has young participants with distributed ages compared to the older population in the validation database. Overall, females in both the study cohorts are ∼57%.

**Table 1.** Comparison of baseline characteristics of study participants in the derivation cohort stratified by the deep neural network model–based phenogroups of diastolic dysfunction.

| Clinical Variable | Overall (n=1252) | Low-Risk (n=588) | High-Risk (n=664) | p-value |
| --- | --- | --- | --- | --- |
| Age, yrs | 62±17 | 54±15 | 69±15 | <0.0001* |
| Males | 539 (43.1%) | 217 (36.9%) | 322 (48.5%) | <0.0001* |
| BMI, kg/m<sup>2</sup> | 29.06±7.42 | 29.42±7.99 | 28.75±6.87 | 0.785 |
| Smoker | 574 (45.8%) | 250 (42.5%) | 324 (48.8%) | 0.030* |
| Diabetes mellitus | 275 (22.0%) | 78 (13.3%) | 197 (29.7%) | <0.0001* |
| Hypertension | 716 (57.2%) | 268 (45.6%) | 448 (67.5%) | <0.0001* |
| Hyperlipidemia | 653 (52.2%) | 270 (45.9%) | 383 (57.7%) | <0.0001* |
| CKD | 266 (21.2%) | 52 (8.8%) | 214 (32.2%) | <0.0001* |
| EF, % | 48.58±25.26 | 51.60±24.74 | 45.91±25.42 | <0.0001* |
| LVMi, g/m<sup>2</sup> | 137.52±168.20 | 118.48±159.81 | 154.39±173.69 | <0.0001* |
| LAVi, mL/m<sup>2</sup> | 44.11±28.39 | 33.20±17.53 | 53.77±32.39 | <0.0001* |
| E, cm/s | 0.88±0.41 | 0.82±0.31 | 0.93±0.48 | <0.0001* |
| A, cm/s | 1.93±2.61 | 2.15±3.24 | 1.73±1.87 | <0.0001* |
| E/A ratio | 1.07±0.56 | 1.03±0.39 | 1.10±0.67 | 0.067 |
| E/e' ratio | 12.00±9.10 | 7.87±3.59 | 15.65±10.78 | <0.0001* |
| TRV, m/s | 8.70±15.17 | 6.43±10.16 | 10.71±18.27 | <0.0001* |
| e', cm/s | 8.11±5.23 | 8.54±2.33 | 7.73±6.82 | <0.0001* |

Values are n (%) or mean ± SD. \*p < 0.05 comparing the high-risk and low-risk groups using the Mann-Whitney U test, where the mean is reported and using the chi-square or Fisher exact test where the frequency is reported.
EF – ejection fraction, LVMi – left ventricular mass index, LAVi – left atrial volume index, E - early mitral inflow velocity, A - late mitral inflow velocity, e' – early diastolic mitral annular velocity, TRV – tricuspid regurgitation velocity, CKD – chronic kidney disease, BMI – body mass index.

**Table 2.** Comparison of baseline characteristics of study participants in the external validation cohort stratified by the deep neural network model–based phenogroups of diastolic dysfunction.

| **Clinical Variable** | **Overall (n=5596)** | **Low-Risk (n=3635)** | **High-Risk (n=1961)** | **p-value** |
| --- | --- | --- | --- | --- |
| Age, yrs | 76 ± 5 | 75 ± 5 | 77 ± 5 | <0.0001* |
| Males | 2409 (43.0%) | 1464 (40.3%) | 945 (48.2%) | <0.0001* |
| BMI, kg/m<sup>2</sup> | 28.49 ± 6.01 | 28.03 ± 5.74 | 29.35 ± 6.41 | <0.0001* |
| Smoker | 317 (5.7%) | 210 (5.8%) | 107 (5.5%) | 0.664 |
| Diabetes mellitus | 1607 (28.7%) | 888 (24.4%) | 719 (36.7%) | <0.0001* |
| Hypertension | 3872 (69.2%) | 2323 (63.9%) | 1549 (79.0%) | <0.0001* |
| Hyperlipidaemia | 2786 (49.8%) | 1962 (54.0%) | 824 (42.0%) | <0.0001* |
| CKD | 1141 (20.4%) | 629 (17.3%) | 512 (26.1%) | <0.0001* |
| EF, % | 65.02 ± 6.78 | 66.95 ± 4.90 | 61.45 ± 8.19 | <0.0001* |
| E, cm/s | 0.68 ± 0.19 | 0.66 ± 0.16 | 0.71 ± 0.23 | <0.0001* |
| A, cm/s | 0.80 ± 0.19 | 0.78 ± 0.17 | 0.85 ± 0.23 | <0.0001* |
| E/A ratio | 0.88 ± 0.30 | 0.87 ± 0.26 | 0.89 ± 0.36 | 0.004* |
| e', cm/s | 5.68 ± 1.48 | 6.15 ± 1.42 | 4.80 ± 1.15 | <0.0001* |
| E/e' ratio | 10.28 ± 4.05 | 9.08 ± 2.67 | 12.51 ± 5.10 | <0.0001* |
| TRV, m/s | 2.38 ± 0.24 | 2.34 ± 0.20 | 2.46 ± 0.28 | <0.0001* |
| LVMi, g/m<sup>2</sup> | 80.24 ± 21.06 | 71.79 ± 13.28 | 95.91 ± 23.70 | <0.0001* |
| LAVi, mL/m<sup>2</sup> | 26.30 ± 9.28 | 23.40 ± 6.33 | 31.68 ± 11.27 | <0.0001* |

Values are n (%) or mean ± SD. \*p < 0.05 comparing the high-risk and low-risk groups using the Mann-Whitney U test, where the mean is reported, and using the chi-square or Fisher exact test where the frequency is reported. EF – ejection fraction, LVMi – left ventricular mass index, LAVi – left atrial volume index, E - early mitral inflow velocity, A - late mitral inflow velocity, e' – early diastolic mitral annular velocity, TRV – tricuspid regurgitation velocity, CKD – chronic kidney disease, BMI – body mass index.

### Development and validation of the emulator model and comparison with the DeepNN model

To mimic the characteristics of the DeepNN model, the proposed emulator model was trained using the same training dataset (n=1009) and validated on the internal validation dataset (n=242) as described previously ^9^. During the training phase, the emulator model showed an accuracy of 97% and an F1 score of 98%. A similar performance is demonstrated by the model in the internal validation dataset (n=242), with an accuracy of 97.12% and an F1-score of 97.45% for predicting the low-risk and high-risk phenogroups of diastolic dysfunction. The comparative results between the previous DeepNN model and the proposed model are shown in Table S1 and Table S2 under section B of the supplemental material. The results indicate that both models exhibited a comparable pattern in predicting low-risk and high-risk phenogroups, with the emulator model outperforming the DeepNN model by producing fewer false positives and negatives.

### Predicting age as a latent variable from the neural activations

To explore and understand the latent space of the echocardiography-driven emulator model, we performed a regression analysis between the age-predicted from neural activations and the actual age. Additionally, we combined the internal and external validation cohorts (n=5529) to conduct a batch prediction and determine if the model’s observations on the younger internal cohort hold for the older population. Figure 2 shows the relationship between the actual age and the predicted biological age of patients in the combined dataset.

**Figure 2.**
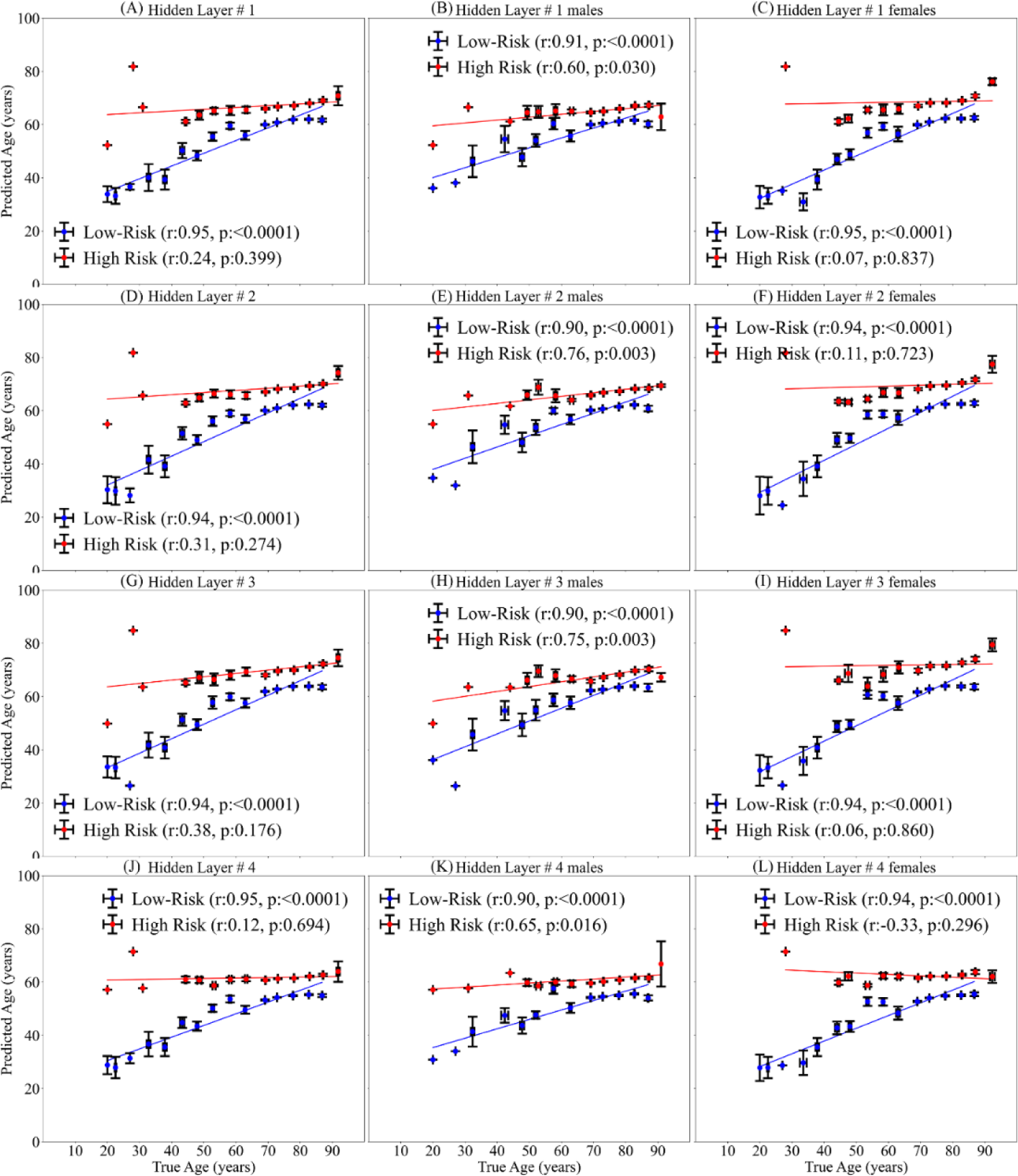
Association between the true age and the biological age predicted using neural activations of the emulator model in overall population (first column), in males (second column), and in females (third column). Four rows correspond to the four hidden layers of the proposed model.

Since the emulator model has four hidden layers, four different sets of neural activations are employed to predict biological age using independent regression models. This association of actual age with the predicted biological age from four hidden layers is illustrated as four rows in Figure 2. Moreover, the association is also indicated separately for an entire population (first column), in males (second column), and females (third column). Figure 2 shows that the correlation coefficient between the actual age and the predicted biological age is higher for the low-risk phenogroup of patients than the high-risk phenogroup of patients. This is consistently true for males and females in all four layers of the deep neural network model. Layer 2 of the model exhibited slightly higher correlation coefficients than other layers for patients’ high-risk phenogroup. For high-risk phenogroups, the correlation coefficients were 0.31 (p=0.274) for the entire population, 0.76 (p<0.01) for males, and 0.11 (p=0.723) for females, respectively. In low-risk phenogroups, these correlation coefficients were 0.94 (p=<0.001) for the entire population, 0.90 (p=<0.001) for males, and 0.94 (p=<0.001) for females, respectively.

Another observation from Figure 2 is that for younger high-risk phenogroup patients, the predicted biological age is higher than their actual age. This overestimation of biological age is possibly due to their elevated values of the echocardiographic variables as discussed in Table 3 to Table 6 in the next section, which govern the neural responses within the network. In contrast, the predicted age for the low-risk phenogroup of patients gradually increased as their actual age advanced, leading to a reduced disparity between the predicted ages of older patients in both the low-risk and high-risk groups. These findings indicate that the emulator model preserves the well-known age-related association with the echocardiographic variables while also being able to discriminate between the low and high-risk phenogroups of patients.

**Table 3.** Analysing the distribution of clinical variables in patients clustered by neural activations of hidden layer 1.

| Variables | C1<br>(n=2681) | C2<br>(n=151) | C3<br>(n=714) | C4<br>(n=1983) |
| --- | --- | --- | --- | --- |
| Age, yrs | 73.83 ± 6.87 | 76.45 ± 9.45 | 77.40 ± 5.94 | 75.49 ± 5.88 |
| E, cm/s | 0.67 ± 0.16 | 0.90 ± 0.32 | 0.77 ± 0.22 | 0.65 ± 0.19 |
| A, cm/s | 0.75 ± 0.20 | 0.61 ± 0.44 | 1.07 ± 0.20 | 0.70 ± 0.22 |
| E/A ratio | 0.86 ± 0.31 | 1.12 ± 0.97 | 0.71 ± 0.19 | 0.84 ± 0.37 |
| EF, % | 67.60 ± 4.85 | 47.77 ± 13.76 | 64.89 ± 7.70 | 62.63 ± 5.68 |
| e', cm/s | 6.56 ± 1.54 | 4.47 ± 1.46 | 4.39 ± 0.92 | 5.19 ± 1.06 |
| E/e' ratio | 8.90 ± 2.55 | 17.28 ± 8.70 | 15.15 ± 5.03 | 10.12 ± 3.20 |
| TRV, m/s | 1.40 ± 1.16 | 2.06 ± 1.20 | 1.51 ± 1.24 | 1.44 ± 1.21 |
| LAVi, mL/m<sup>2</sup> | 23.18 ± 6.29 | 48.92 ± 23.32 | 28.72 ± 8.69 | 28.94 ± 9.09 |
| LVMi, g/m<sup>2</sup> | 69.32 ± 12.66 | 135.67 ± 38.01 | 87.74 ± 20.90 | 88.27 ± 17.67 |

EF – ejection fraction, LVMi – left ventricular mass index, LAVi – left atrial volume index, E - early mitral inflow velocity, A - late mitral inflow velocity, e' – early diastolic mitral annular velocity, TRV – tricuspid regurgitation velocity, CKD – chronic kidney disease, BMI – body mass index.

**Table 4.** Analysing the distribution of clinical variables in patients clustered by neural activations of hidden layer 2.

| Variables | C1<br>(n=171) | C2<br>(n=2682) | C3<br>(n=402) | C4<br>(n=2274) |
| --- | --- | --- | --- | --- |
| Age, yrs | 76.56 ± 8.83 | 73.70 ± 6.81 | 77.05 ± 6.79 | 75.96 ± 5.83 |
| E, cm/s | 0.88 ± 0.32 | 0.66 ± 0.16 | 0.82 ± 0.26 | 0.66 ± 0.18 |
| A, cm/s | 0.67 ± 0.46 | 0.73 ± 0.20 | 0.91 ± 0.39 | 0.79 ± 0.24 |
| E/A ratio | 0.95 ± 0.78 | 0.87 ± 0.30 | 0.84 ± 0.57 | 0.80 ± 0.32 |
| EF, % | 47.61 ± 13.20 | 67.35 ± 4.87 | 60.69 ± 8.75 | 64.12 ± 5.64 |
| e', cm/s | 4.22 ± 1.24 | 6.60 ± 1.50 | 4.47 ± 1.19 | 5.05 ± 1.06 |
| E/e' ratio | 17.59 ± 8.98 | 8.70 ± 2.46 | 15.82 ± 5.41 | 10.85 ± 3.38 |
| TRV, m/s | 1.92 ± 1.25 | 1.41 ± 1.16 | 1.69 ± 1.24 | 1.42 ± 1.21 |
| LAVi, mL/m<sup>2</sup> | 45.99 ± 22.30 | 23.33 ± 6.39 | 35.48 ± 11.93 | 27.58 ± 8.07 |
| LVMi, g/m<sup>2</sup> | 133.60 ± 36.26 | 69.60 ± 12.73 | 101.05 ± 22.41 | 85.26 ± 16.87 |

EF – ejection fraction, LVMi – left ventricular mass index, LAVi – left atrial volume index, E - early mitral inflow velocity, A - late mitral inflow velocity, e' – early diastolic mitral annular velocity, TRV – tricuspid regurgitation velocity, CKD – chronic kidney disease, BMI – body mass index.

**Table 5.** Analysing the distribution of clinical variables in patients clustered by neural activations of hidden layer 3.

| Variables | C1<br>(n=1866) | C2<br>(n=2654) | C3<br>(n=138) | C4<br>(n=871) |
| --- | --- | --- | --- | --- |
| Age, yrs | 75.83 ± 5.65 | 73.66 ± 6.83 | 76.57 ± 9.00 | 76.78 ± 6.69 |
| E, cm/s | 0.65 ± 0.17 | 0.66 ± 0.16 | 0.90 ± 0.34 | 0.76 ± 0.23 |
| A, cm/s | 0.79 ± 0.23 | 0.73 ± 0.20 | 0.72 ± 0.47 | 0.85 ± 0.35 |
| E/A ratio | 0.80 ± 0.30 | 0.88 ± 0.31 | 1.00 ± 0.79 | 0.80 ± 0.48 |
| EF, % | 64.62 ± 5.33 | 67.35 ± 4.86 | 47.87 ± 13.88 | 60.89 ± 8.27 |
| e', cm/s | 5.14 ± 1.04 | 6.61 ± 1.49 | 4.10 ± 1.21 | 4.57 ± 1.13 |
| E/e' ratio | 10.44 ± 3.09 | 8.67 ± 2.45 | 18.71 ± 9.17 | 14.08 ± 5.03 |
| TRV, m/s | 1.39 ± 1.20 | 1.41 ± 1.16 | 1.96 ± 1.26 | 1.61 ± 1.24 |
| LAVi, mL/m<sup>2</sup> | 26.91 ± 7.63 | 23.37 ± 6.41 | 47.72 ± 23.97 | 32.84 ± 11.05 |
| LVMi, g/m<sup>2</sup> | 83.15 ± 15.91 | 69.69 ± 12.77 | 138.15 ± 37.18 | 97.41 ± 21.53 |

EF – ejection fraction, LVMi – left ventricular mass index, LAVi – left atrial volume index, E - early mitral inflow velocity, A - late mitral inflow velocity, e' – early diastolic mitral annular velocity, TRV – tricuspid regurgitation velocity, CKD – chronic kidney disease, BMI – body mass index.

**Table 6.** Analysing the distribution of clinical variables in patients clustered by neural activations of hidden layer 4.

| Variables | C1<br>(n=4466) | C2<br>(n=110) | C3<br>(n=584) | C4<br>(n=369) |
| --- | --- | --- | --- | --- |
| AGE, yrs | 74.54 ± 6.45 | 76.48 ± 9.11 | 76.55 ± 6.45 | 77.07 ± 7.17 |
| E, cm/s | 0.66 ± 0.17 | 0.90 ± 0.33 | 0.72 ± 0.21 | 0.83 ± 0.26 |
| A, cm/s | 0.75 ± 0.21 | 0.79 ± 0.47 | 0.86 ± 0.30 | 0.82 ± 0.42 |
| E/A ratio | 0.85 ± 0.31 | 1.00 ± 0.76 | 0.77 ± 0.36 | 0.85 ± 0.63 |
| EF, % | 66.27 ± 5.19 | 46.48 ± 14.41 | 62.56 ± 6.36 | 57.92 ± 10.18 |
| e', cm/s | 6.02 ± 1.51 | 3.94 ± 1.08 | 4.63 ± 1.08 | 4.49 ± 1.20 |
| E/e' ratio | 9.37 ± 2.85 | 20.00 ± 9.65 | 12.95 ± 4.07 | 15.55 ± 5.73 |
| TRV, m/s | 1.40 ± 1.17 | 1.96 ± 1.24 | 1.55 ± 1.23 | 1.71 ± 1.25 |
| LAVi, mL/m<sup>2</sup> | 24.80 ± 7.15 | 47.42 ± 24.89 | 30.10 ± 9.31 | 37.57 ± 13.34 |
| LVMi, g/m<sup>2</sup> | 75.11 ± 15.55 | 138.90 ± 39.97 | 92.34 ± 19.56 | 106.75 ± 23.91 |

EF – ejection fraction, LVMi – left ventricular mass index, LAVi – left atrial volume index, E - early mitral inflow velocity, A - late mitral inflow velocity, e' – early diastolic mitral annular velocity, TRV – tricuspid regurgitation velocity, CKD – chronic kidney disease, BMI – body mass index.

### Examining patient pathways through the network and the impact of echocardiographic variables

To further answer the question of how the echocardiographic variables of patients are travelling through the neural network, we dissected the neural network activations. We performed hierarchical clustering for all patients ^20^. The details of hierarchical clustering are provided in supplemental material (section C). Hierarchical clustering separated the patients into four row-wise clusters (illustrated in Figure 3), such as C1 to C4, whose clinical variables in each of the four layers are shown in Tables 3 to 6, respectively. Table 3 to Table 6 present the clustering results, revealing that out of four clusters, the high-risk phenogroup of patients predominantly falls into clusters C2, C1, C3, and C2, respectively, across four hidden layers.

**Figure 3.**
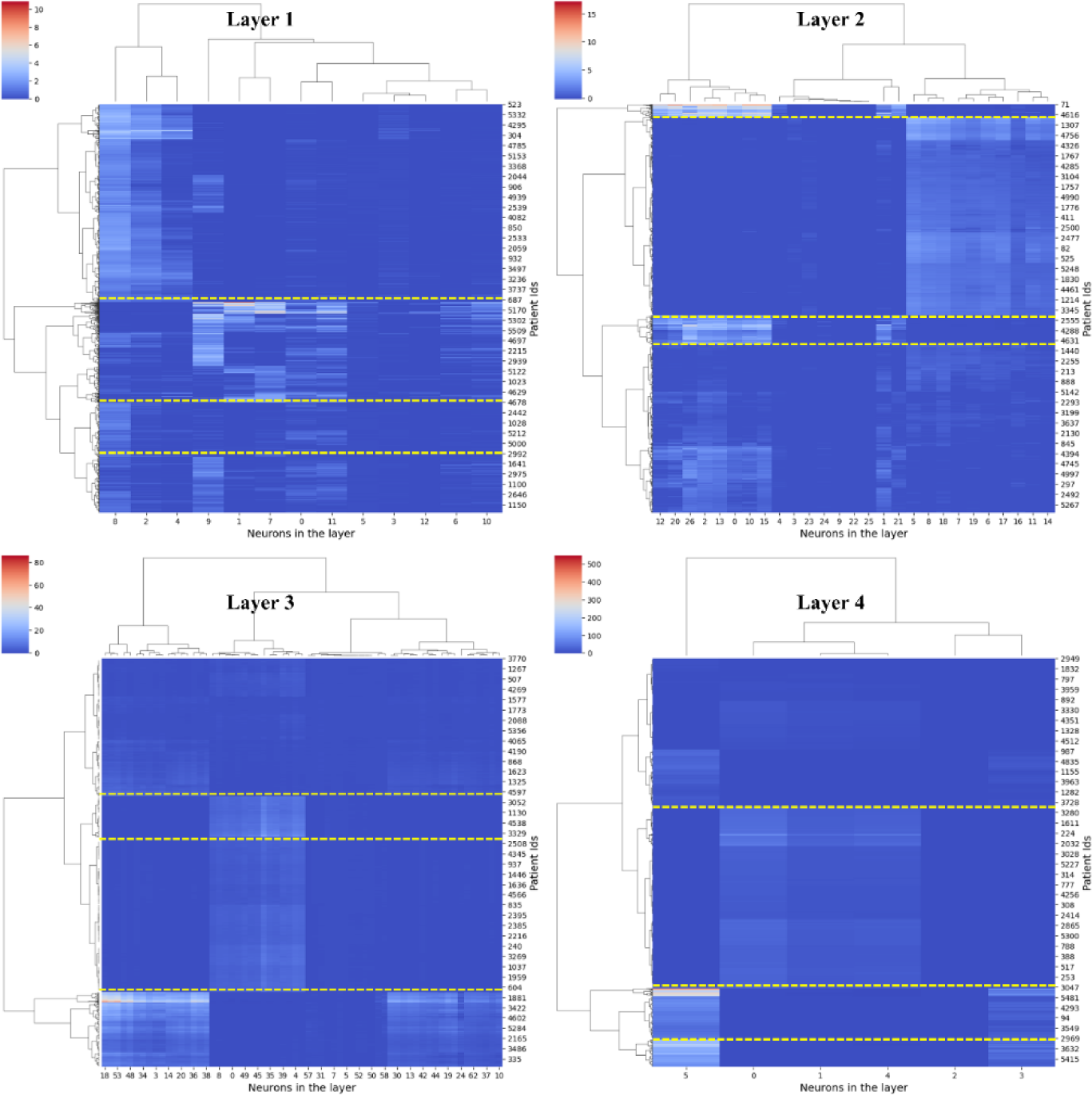
Hierarchical clustering of the neural network activations indicating four unique sets of clusters of patients in each hidden layer.

Additionally, it is worth noting that apart from clustering patients, neurons exhibiting similar activations have also been grouped. This observation might suggest that these neurons play a role in the distinct pathways taken by low-risk or high-risk phenogroups of patients. In our investigation of patient pathways across the four layers of the network, we identified the three most common neural pathways followed by the high-risk and low-risk phenogroups of patients. For the high-risk phenogroups, the three distinct neural pathways are 10-27-16-6, 10-3-54-6, and 8-3-54-6, while for the low-risk patients, the pathways are 9-6-23-1, 9-6-5-1, and 9-6-40-1. Figure 4 depicts the pathways followed by three sample patients from the low-risk and high-risk phenogroups, showcasing the distinct neural trajectories identified in our analysis.

**Figure 4.**
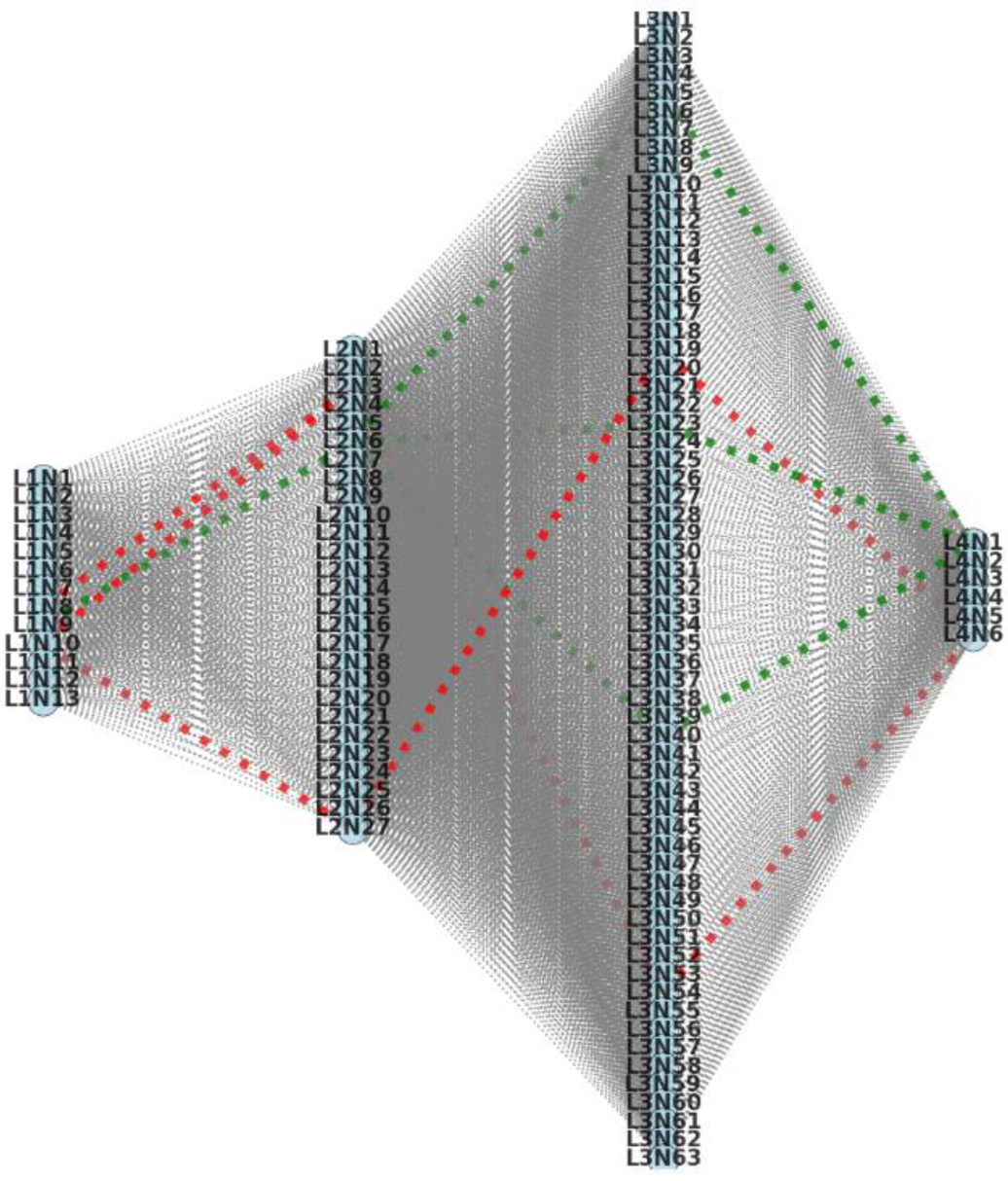
Neural network pathways followed by three sample patients from low-risk and high-risk phenogroups. L1-L4 are four hidden layers. <u>High-risk patient P1</u> (pathway L1N10-L2N27-L3N19-L4N6): EF=43.6, E=0.65, A=1.18, E/A=0.6, e’=2.9, E/e’= 20.6, TRV= 2.76, LAVi= 25.31, LVMi=106, <u>high-risk patient P2</u> (pathway L1N10-L2N3-L3N54-L4N6): EF=57.6, E=0.47, A=0.91, E/A=0.5, e’=5.7, E/e’= 7.9, TRV= 2.53, LAVi= 20.9, LVMi=94.32, <u>high-risk patient P3</u> (pathway L1N8-L2N3-L3N54-L4N6): EF=40.6, E=0.38, A=0.69, E/A=0.5, e’=6.5, E/e’= 4.4, TRV= 2.42, LAVi= 31.46, LVMi=122.32. <u>Low-risk patient P1</u> (pathway L1N9-L2N6-L3N23-L4N1): EF=82.2, E=0.85, A=0.82, E/A=1, e’=5.7, E/e’= 9.7, TRV= 1.98, LAVi= 20.74, LVMi=83.55, <u>low-risk patient P2</u> (pathway L1N9-L2N6-L3N5-L4N1): EF=75.6, E=0.54, A=0.89, E/A=0.6, e’=4.5, E/e’= 8.2, TRV= 2, LAVi= 19.99, LVMi=76.23, low-risk patient P3 (pathway L1N9-L2N6-L3N40-L4N1): EF=68.4, E=0.84, A=0.81, E/A=1, e’=5.2, E/e’= 10.7, TRV= 2.46, LAVi= 14.56, LVMi=106.54.

### Prediction of sex as a latent variable from the neural activations

To further investigate the latent space of the emulator model, five ML algorithms are trained to predict sex as a latent variable from the neural activations of a deep neural network. Table 5 lists the six performance evaluation metrics for five algorithms for internal and external validation cohorts. Across all ML-based algorithms for predicting sex, the average accuracy was 60% and 59%, respectively (Table 7), in the internal and external validation cohorts. Logistic regression algorithms reported better classification performance with an AUC of 0.65 in both validation cohorts.

**Table 7.** Predicting sex as a latent variable from the second layer of neural network model.

| Database | Classifier | Sensitivity (%) | Specificity (%) | F-Score (%) | ACC (%) | AUC |
| --- | --- | --- | --- | --- | --- | --- |
| Interval validation cohort (n=243) | SVM | 41.90 | 70.29 | 46.32 | 58.02 | 0.61 |
|  | LR | 87.62 | 35.51 | 64.34 | 58.02 | 0.65 |
|  | Bagging | 46.67 | 74.64 | 51.85 | 62.55 | 0.63 |
|  | RF | 44.76 | 70.29 | 48.70 | 59.26 | 0.64 |
|  | XGBoost | 51.43 | 70.29 | 54.00 | 62.14 | 0.62 |
|  | Mean | 54.48 | 64.20 | 53.04 | **60.00** | 0.63 |
| External validation cohort (n=5596) | SVM | 27.07 | 88.14 | 37.92 | 61.85 | 0.62 |
|  | LR | 87.46 | 25.67 | 61.21 | 52.27 | 0.65 |
|  | Bagging | 38.77 | 78.07 | 46.21 | 61.15 | 0.62 |
|  | RF | 36.20 | 79.29 | 44.25 | 60.74 | 0.62 |
|  | XGBoost | 41.59 | 72.23 | 46.65 | 59.04 | 0.61 |
|  | Mean | 46.22 | 68.68 | 47.25 | **59.01** | 0.62 |

Although the accuracy and AUC values are less, it should be noted that this was the first attempt to show the association between the hidden layer neural activations and sex.

## Discussion

Diving into the hidden representations of deep neural networks is always an interesting yet challenging task. Our previous study shows that the echocardiographic deep neural network (i.e., DeepNN) can predict the risk of LVDD from the nine diastolic function parameters in younger and elderly cohorts ^9 11^. In this study, we attempted to dissect the model’s internal architecture to investigate latent space’s association with the two demographic parameters, such as age and sex. The network dissection revealed that the hidden layers of the emulator model, although trained to predict the LVDD, exhibit an association with the actual age of patients, validating our hypothesis for this study. Although accuracy for predicting sex from the latent space of the emulator model is low, we still demonstrated the capability of the hidden neural activations to provide some information about the sex of patients.

We know that the biological age differs from the chronological actual age and involves the time elapsed and several biological and physiological factors, including genetics, nutrition, and comorbidities ^21^. Therefore, it becomes essential to understand how the structural changes in the heart, captured by the echocardiography-driven neural network, can be used to predict the biological age. The present study results show that the low-risk and high phenogroups of LVDD are associated with actual age, and this association can also be preserved by the neural network indicating a similar association between the phenogroups and the echocardiography-derived biological age. That means by using nine echocardiography parameters, the emulator model can distinguish between low-risk and high-risk phylogroups, and by using echocardiography-driven biological age predicted from the latent space of the model, a similar distinction can also be possible.

In this study, we included participants from two different cohorts. The derivation cohort consisted of participants who were younger, middle-aged, and older people with ages ranging from 18 years to 97 years. The majority of participants in the derivation cohort were young or middle-aged. In contrast to the derivation cohort, the external validation cohort included participants between the ages of 66 and 90 years old, considered to be older members of the cohort. Our findings have shown that hidden neural activations from the proposed emulator model in both cohorts were capable of providing an idea about the age and sex of patients. However, for the younger age group, the hidden neural activations showed an overestimated age; for older adults, the predicted age was underestimated compared to the actual age of patients. This could be due to the dissimilarities in actual age and biological age (or “heart age”) captured by the echocardiography parameters. The age information captured by the hidden neural activations reflects the biological age of patients. From a previous study by Okura and colleagues ^22^, it is known that aging leads to structural changes in the heart that may change a person’s biological age. The overestimation and underestimation of biological age concerning actual age show that the model can capture the changes in the heart’s structure. Our study results (Figure 2) further point to the ability of the hidden neural activations of the emulator model to capture these changes in high-risk phenogroups in a much more pronounced manner. We have also observed that the information captured by neural activations in the high-risk group significantly differs from the low-risk LVDD phenogroups in both sexes (Figure 2).

Previous attempts were made to predict the actual age and sex from the echocardiograms ^22 23^ and electrocardiogram (ECG) ^24^, however, the age-dependent LVDD severity prediction was not investigated. Okura et al. ^22^ showed that aging leads to a more pronounced deterioration of diastolic function in females than in males ^22^. Also, the incidence of diastolic heart failure is higher in elderly females than in males ^22^. The authors also demonstrated a significant association of actual age and sex with the left ventricular function ^22^, which may further change LVDD severity. Although the study by Okura et al. ^22^ made the observations considering the chronological age, it would have been more interesting to investigate similar observations with echocardiography-driven biological age. Another study by Attia et al. ^24^ predicted the biological age and sex from the 12-lead ECG signals using the convolutional neural network-based model. The authors showed that ECG signals have the potential to predict the biological age and sex of patients. However, it was a direct approach with age and sex being used as labels to train the CNN model, and no latent information within hidden neural activations was explored.

This study on the dissection of the deep neural network models points to a vital aspect of echocardiography-driven AI models that they can capture biological age or sex information, even if they are not primarily trained to do so. This study is a first-ever attempt to dissect the latent space of a deep neural network for providing the age and sex-related association with the LVDD phenogroups. Further research should include other comorbidities when predicting LVDD phenotypes and then investigate how latent space captures the effect of comorbidities when predicting actual age.

## Conclusion

This study proposed an echocardiographic deep neural network-based emulator that characterizes patients with low or high potential burden of LVDD. The dissection of the echocardiographic emulator can provide additional information on the biological age governed by the latent space of the deep neural network. The echocardiography-driven biological age can help in distinguishing patients with LVDD. Our findings suggest that the neural network-based architectures can preserve the well-known association between echocardiographic parameters and the actual age of patients while being able to discriminate the potential burden of LVDD in both younger and older populations. Therefore, this study could be helpful to provide an age-agnostic solution that can assist clinicians with new pathophysiological insights.

## Data Availability

All data produced in the present study are available upon reasonable request to the authors

## Acknowledgments

Authors would like to thank Dr. Quincy Hathaway, MD, PhD from West Virginia University School of Medicine, WV, USA, for providing valuable feedback on this manuscript that significantly improved the quality and reliability of this manuscript.

