## Supplemental for "Analyzing the Mechanism Behind Age-agnostic Prediction of Diastolic Dysfunction Using Echocardiography Variables in Deep Neural Networks"

**Supplementary Material**

### A. Echocardiographic parameters

1. *Left Ventricular Ejection Fraction*: Left ventricular ejection fraction (LVEF) is a measure that tells us how well the left side of the heart is pumping blood out to the body. It is expressed as a percentage and represents the proportion of blood that is pumped out of the left ventricle with each heartbeat. A normal LVEF is typically around 50-70%. A lower LVEF indicates that the heart is not pumping blood as effectively as it should be. This can be a sign of a heart condition or damage to the heart muscle. A reduced LVEF can lead to symptoms like fatigue, shortness of breath, and fluid retention. LVEF is an important measure used by doctors to assess heart function and determine appropriate treatment options for patients with heart problems.
2. *Early and late diastolic transmitral flow velocities*: "E" and "A" refer to the two peak velocities observed in the transmitral flow pattern during the cardiac cycle. These velocities are obtained using pulsed-wave Doppler imaging and provide information about the filling of the left ventricle. "E" represents the early diastolic transmitral flow velocity. It corresponds to the speed at which blood flows from the left atrium to the left ventricle during early diastole, which is the resting phase of the cardiac cycle when the ventricles relax and fill with blood. The E velocity is influenced by factors such as left ventricular relaxation and compliance. "A" represents the late diastolic transmitral flow velocity. It represents the speed at which blood flows from the left atrium to the left ventricle during late diastole, just before the next contraction. The A velocity is influenced by factors such as left atrial contraction and left ventricular compliance. The E/A ratio, which is calculated by dividing the E velocity by the A velocity, is an important parameter used to assess left ventricular diastolic function. An abnormal E/A

ratio can indicate diastolic dysfunction, where there is impaired relaxation or abnormalities in the filling of the left ventricle. The E and A velocities, along with the E/A ratio, help evaluate the filling dynamics of the left ventricle and provide valuable information about cardiac function, especially in the context of diastolic function and heart failure.

3. *Early diastolic mitral annular velocity (e')*: It is a specific measurement obtained through tissue Doppler imaging during echocardiography. It provides information about the movement and velocity of the mitral valve annulus, which is a structure near the mitral valve, during early diastole. e' represents the speed at which the mitral annulus moves towards the apex of the heart during early diastole, which is the resting phase when the ventricles relax and fill with blood. It reflects the rate of relaxation and compliance of the left ventricle. The e' velocity measurement is useful for assessing left ventricular diastolic function. Abnormal E' values can indicate diastolic dysfunction, where the left ventricle does not relax adequately or there are abnormalities in the filling of the ventricle. Changes in E' velocity can also provide insights into the overall cardiac function and help in the diagnosis and management of various heart conditions.
4. *Left ventricular mass index*: The left ventricular mass index (LVMI) is a measurement used in cardiology to assess the size of the left ventricle, the main pumping chamber of the heart. It is calculated by dividing the left ventricular mass by the body surface area. The left ventricular mass is the weight or mass of the muscular wall of the left ventricle. By dividing it by the body surface area, which takes into account a person's height and weight, the LVMI adjusts for body size and allows for a standardized comparison across individuals. The LVMI measurement provides valuable information about the size and thickness of the left ventricle. An increased LVMI is often associated with conditions such as hypertension (high blood pressure), hypertrophic cardiomyopathy (thickened

heart muscle), or other cardiac abnormalities. Monitoring the LVMI can help assess changes in the size and mass of the left ventricle over time and guide treatment decisions for various heart conditions. It is typically obtained through imaging techniques such as echocardiography or cardiac MRI.

5. *E/e' ratio*: The E/e' ratio is a calculation used in echocardiography to assess left ventricular filling pressures. It combines two measurements: the early diastolic transmitral flow velocity (E) and the early diastolic mitral annular velocity (e'). An elevated E/e' ratio suggests increased filling pressures, which can be associated with conditions of diastolic dysfunction.
6. *Tricuspid regurgitation velocity*: Tricuspid regurgitation velocity refers to the speed at which blood flows in the opposite direction across the tricuspid valve during diastole. It is measured using Doppler echocardiography. Tricuspid regurgitation occurs when the tricuspid valve, located between the right atrium and right ventricle, does not close properly, allowing blood to leak back into the right atrium during ventricular relaxation. The regurgitant flow generates a velocity that can be measured using Doppler ultrasound.

**B. Establishing equivalence between the DeepNN model and the emulator model.**

**Table S1. The performance evaluation metrics comparing DeepNN and Emulator models.**

| PE Metric | DeepNN | Emulator |
|---|---|---|
| Sensitivity, (%) | 100.00 | 100.00 |
| Specificity, (%) | 93.58 | 97.25 |
| Kappa | 0.94 | 0.97 |
| F1-Score, (%) | 97.45 | 98.89 |
| Accuracy, (%) | 97.12 | 98.77 |
| AUC | 0.998 | 0.993 |

**Table S2. Confusion matrix for DeepNN and Emulator models.**

| | Risk Category | DeepNN | | Emulator | |
|---|---|---|---|---|---|
| | | Low-Risk | High-Risk | Low-Risk | High-Risk |
| True Labels | Low-Risk | 102 | 9 | 106 | 3 |
| | High-Risk | 0 | 134 | 0 | 134 |
| | | Predicted Labels | | | |

### C. Hierarchical clustering

Hierarchical clustering is a widely utilized technique in data analysis and pattern recognition that aims to group similar data points into clusters based on their similarities [27]. It constructs a hierarchical structure called a dendrogram, which represents the relationships between the data points. The process begins by considering each data point as an individual cluster and subsequently merging clusters based on their similarity, gradually forming the hierarchy. Hierarchical clustering offers several advantages, including its ability to uncover the inherent hierarchical structure within the data and its suitability for visualization using dendrograms. It does not require predefining the number of clusters, making it a flexible approach. Furthermore, it can accommodate various data types and distance metrics, allowing customization to suit the specific problem at hand. In this study, we are using Agglomerative clustering, also known as bottom-up clustering, which starts by treating each data point as a separate cluster and iteratively merging them until a single cluster is formed. We used the *cluster map* functionality of the *seaborn* library to generate the hierarchical clustering of the neural activations in the hidden layers.
